## Supplemental Table 1 for "Top-funded digital health companies offering lifestyle interventions for dementia prevention: Company overview and evidence analysis"

### Supplementary Table 1:

| Search category | Search keywords |
|---|---|
| **Verticals, methods & industries** | (Monitoring Equipment OR diagnostic OR HealthTech OR healthcare devices OR connected health* OR Therapeutic Devices OR Digital Health OR digital health* OR health* technology OR health* app* OR wearables OR Mobile health OR mhealth OR digital therapeutics OR mobile app OR personal health OR virtual care OR e-health OR assistive technology OR telehealth OR telemedicine OR health* platform OR healthcare it OR data management OR Artificial Intelligence & Machine Learning OR Cloud data services OR analytics OR health* diagnostics OR Big Data OR information OR digital OR data OR biometrics OR home health care OR medtech OR self monitoring) |
| **AND** | |
| **Dementia** | dementia OR alzheimer OR memory loss OR cognitive decline OR Alzheimer's disease OR confusion OR impairment OR brain degeneration OR senility OR dementia symptoms OR dementia diagnosis OR dementia treatment OR brain aging OR neurodegenerative disorder OR mental decline OR forgetfulness OR brain function OR brain health OR brain performance |
| **AND** | |
| **Management & prevention** | AND (dementia management OR dementia control OR dementia prevention OR dementia monitoring OR dementia detection OR assessment OR diagnosis OR disease diagnosis OR improvement OR disease monitoring OR disease management OR risk reduction OR disease prevention OR prevention) |
