## Supplemental Table 2 for "Top-funded digital health companies offering lifestyle interventions for dementia prevention: Company overview and evidence analysis"

### Supplementary Table 2:

| Search category | Search keywords |
|---|---|
| **Verticals, methods & industries** | therapeutics, Psychology, Personal Health, Nutrition, mHealth, Medical Device, Medical, Employee Benefits, Elder Care, Dietary Supplements, Assistive Technology, Assisted Living, Alternative Medicine |
| **Dementia and Management & prevention** | dementia control, dementia treatment, dementia symptoms, dementia monitoring, dementia prevention, dementia management, dementia diagnosis, Alzheimer diagnosis, Alzheimer management, Alzheimer prevention, Alzheimer monitoring, Alzheimer symptoms, Alzheimer treatment, Alzheimer control, brain performance, brain health, brain function, brain aging, brain generation, mental decline, neurodegenerative disorder, momory loss, cognitive decline |
